# Polyethylene Glycol-Mediated Directional Conjugation of Biological Molecules for Enhanced Immunoassays at the Point-of-Care

**DOI:** 10.1101/2023.04.22.23288974

**Authors:** Dheerendranath Battalapalli, Purbali Chakraborty, Disha Jain, Stephen K. Obaro, Umut Gurkan, Robert A. Bonomo, Mohamed Draz

**Affiliations:** Department of Medicine, Case Western Reserve University School of Medicine, Cleveland, OH 44106, USA; Research Service, Louis Stokes Cleveland Department of Veterans Affairs Medical Center, Cleveland, OH, USA; University of Nebraska Medical Center, Omaha, NE 68198, USA; Mechanical and Aerospace Engineering Department, Case Western Reserve University, Cleveland, Ohio, USA; Department of Biomedical Engineering, Case Western Reserve University, Cleveland, Ohio, USA; Case Comprehensive Cancer Center, Case Western Reserve University, Cleveland, Ohio, USA; Molecular Biology and Microbiology, Case Western Reserve University School of Medicine, Cleveland, OH, USA; CWRU-Cleveland VAMC Center for Antimicrobial Resistance and Epidemiology (Case VA CARES) Cleveland, OH, USA; Department of Biomedical Engineering, Cleveland Clinic, Cleveland, OH 44106, USA

**Author notes:** Corresponding Author: Mohamed Draz.

**Keywords:** Biological molecules, antibody, directional, site-specific, controlled, conjugation, immunoassay, screen-printed electrodes, point of care

## Abstract

Rapid and reliable point-of-care (POC) diagnostic tests can make a significant impact on global health. One of the most common approaches for developing POC systems is the use of target-specific biomolecules. However, the conjugation of biomolecules can result in decreased activity, which may compromise the analytical performance and accuracy of the developed systems. Here, we present a polymer-based cross-linking protocol for controlled and directed conjugation of biological molecules. Our protocol utilizes a bifunctional thiol-polyethylene glycol (PEG)-hydrazide polymer to enable site-directed conjugation of IgG antibodies to the surface of screen-printed metal electrodes. The metal surface of the electrodes is first modified with thiolated PEG molecules, leaving the hydrazide groups available to react with the aldehyde group in Fc fragments of the oxidized IgG antibodies. Using *Klebsiella pneumoniae* carbapenemase-2 (KPC2) antibody as a model antibody used for antimicrobial resistance (AMR) testing, our results demonstrate a ∼10-fold increase in antibody coupling, compared with the standard *N*-hydroxysuccinimide (NHS)-based conjugation chemistry, and effective capture (> 94%) of the target KPC2 enzyme antigen on the surface of modified electrodes. This straightforward and easy-to-perform strategy of site-directed antibody conjugation can be engineered for coupling other protein and non-protein-based biological molecules commonly used in POC testing and development, thus enhancing the potential for improved diagnostic accuracy and performance.

## Introduction

The demand for point-of-care (POC) medicine is rapidly increasing in our modernized and globalized world. POC diagnostics have revolutionized healthcare by providing rapid and accurate disease diagnosis at the patient’s bedside or in a clinical setting [1]. Immunoassays are widely used in POC diagnostics for detecting analytes such as proteins, nucleic acids, and small molecule [2]. However, the sensitivity and specificity of immunoassays are limited by the availability and orientation of the antibodies on the surface of the assay platform [3]. Conventional conjugation methods involve random and non-specific chemical reactions between the antibody and the conjugation partner, such as *N*-hydroxysuccinimide (NHS) ester or maleimide chemistry, which can result in low yields, heterogeneous products, and reduced antibody activity [4, 5]. Site-specific modification strategies have been developed to overcome these limitations by enabling the selective conjugation of antibodies to specific sites, improving the availability and orientation of the antibodies on the surface of the assay platform [6-13].

Chemical conjugation using functional polymers such as polyethylene glycol (PEG) has emerged as a superior alternative to traditional conjugation methods [14]. PEG can be easily conjugated to antibodies through various chemistries and offers several advantages over other polymers, including high water solubility, low toxicity, and low immunogenicity [15-17]. Moreover, PEG conjugation can enhance the stability and shelf-life of the antibody, increase the solubility and bioavailability of the conjugate, and reduce the clearance rate from the body [18-20]. These properties make PEG an ideal candidate for conjugation with antibodies in POC diagnostics.

One of the major advantages of chemical conjugation using functional polymers like PEG is enabling site-specific modification for antibody conjugation [21-23]. These approaches have gained significant attention in the development of POC diagnostics. They enable the selective and controlled conjugation of antibodies to specific sites, reducing the risk of antibody denaturation and loss of function [24]. Moreover, they offer enhanced performance and accuracy in immunoassays by improving the availability and orientation of the antibodies on the assay platform. Here, we present a novel approach for PEG-mediated site-specific antibody conjugation for the development of a POC-based electrode microchip immunoassay. We demonstrate the efficacy of our approach by developing anti-Klebsiella pneumoniae carbapenemase-2 (KPC2) antibody-modified screen-printed gold (Au) electrodes with improved conjugation and target-capture efficiency compared to conventional conjugation methods. In our approach, a heterobifunctional linker of thiol-PEG-hydrazide (SH-PEG-hydrazide) was used to modify the carbohydrate residues of the Fc region of polyclonal anti-KPC2 IgG antibody. The antibody modified with PEG-thiol is then allowed to react with the surface of Au electrodes via the well-known thiol-metal bonding chemistry. Our approach represents a significant advancement in the development of conjugation chemistry for POC diagnostics, offering improved performance and accuracy for rapid and reliable disease diagnosis.

## Results and Discussion

The successful development of effective POC diagnostics relies on the use of optimized conjugation methods that enable the selective and controlled conjugation of antibodies to the assay platform [25, 26]. This can significantly improve the sensitivity and specificity of immunoassays, enabling accurate and reliable disease diagnosis at the POC [1, 27]. Over the past decade, there has been a significant advancement in engineering PEG chemistry [19, 28]. A majority of current applications for PEG include therapeutic and diagnostic [16, 17]. PEGylation of antibodies, in particular, has been known to increase the half-life and reduce the non-specific binding of antibodies and hence leads to an economical production and usage of immunoassays [29, 30]. Here, we describe a protocol for the site-directed conjugation of a bifunctional 20 kDa PEG linker to the Fc region of polyclonal anti-KPC2-antibody to assist binding to gold screen-printed electrodes (Au-SPEs), one of the most common platforms used for electrochemical-based immunoassays at the POC [31-33]. This resulted in a packed conjugation of antibody molecules on the surface of Au-SPE, which is more favorable as there were more functional moieties to bind the antigen molecule. This further resulted in enhanced capture of KPC2 antibody suggesting its potential for sensitive detection of antimicrobial resistance (AMR) causing carbapenemases enzyme is possible in surface-modified Au-SPEs using an approach of directional conjugation.

To develop screen-printed electrodes for the detection of KPC2 enzyme at POC, we designed a site-directed conjugation chemistry that relies on using a heterobifunctional PEG polymer to link oxidized polyclonal anti KPC2 antibodies to the surface of Au-SPEs [34, 35] (**Figure 1a**). PEGylation of the Au-SPE was done using thiolated-PEG-hydrazide (SH-PEG-hydrazide) which forms a non-covalent gold-thiol bond, leaving the PEG-hydrazide arm free to react [36]. Anti-KPC2 polyclonal antibody was oxidized using sodium metaperiodate to convert the carbohydrate group in glycoproteins to reactive aldehyde groups. The free hydrazide group on the surface of Au-SPE reacted with the aldehyde group on the antibody Fc region for its directional conjugation (**Figure 1a**).

**Figure 1.**
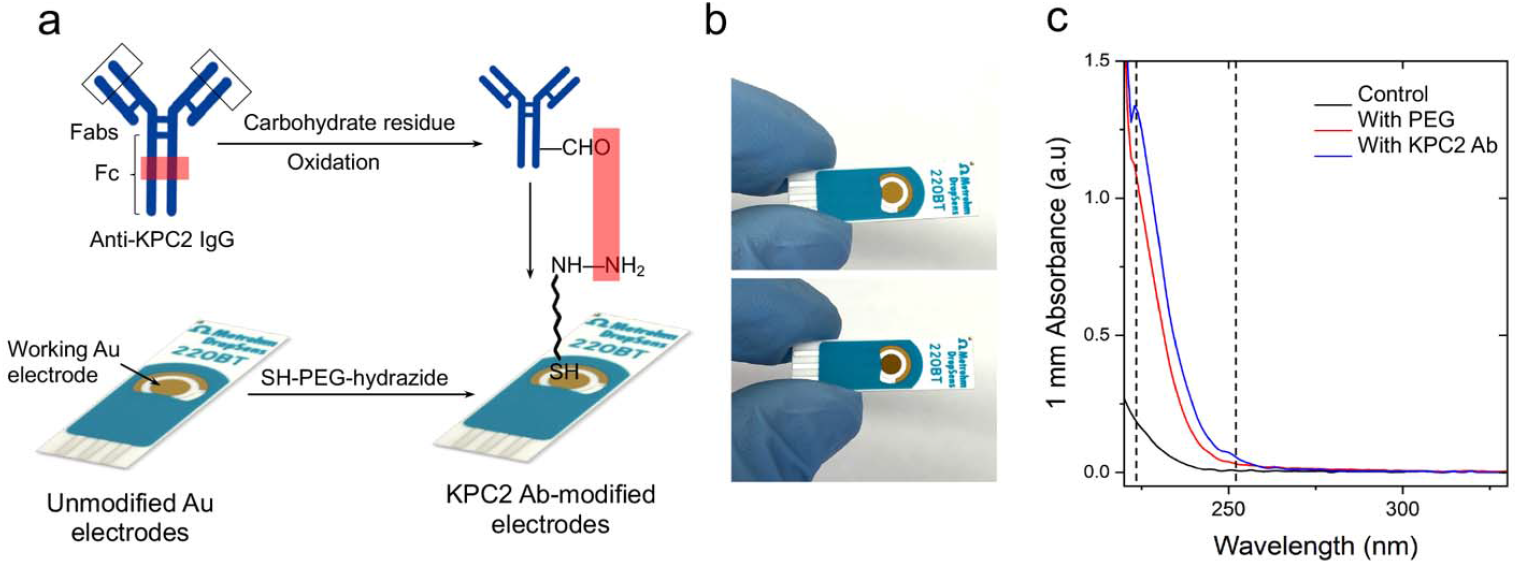
PEG-medicated conjugation of IgG antibodies to the surface of screen-printed gold electrodes. (a) Schematic presentation of the protocol for chemical modification of Au-SPE with SH-PEG-hydrazide followed by coupling with antibody. (b) Digital images of unmodified (top) and modified Au-SPE (bottom), showing a change in color of the surface post-modification. (c) UV-Vis spectrum of control Au-SPE (without any treatment; black line), PEGylated Au-SPE (red line) and antibody-coupled Au-SPE (blue line). The surface of Au-SPEs was digested with TCEP, and the eluted solution was tested using a NanoDrop spectrophotometer to measure the absorbance.

To evaluate the efficiency of the developed PEG-based site-directed conjugation approach, we tested the presence of antibodies on the surface of Au-SPEs using multiple techniques, including colorimetry, fluorometry, Ultraviolet-visible (UV-Vis), and Fourier-transform infrared (FTIR) spectroscopy and sodium dodecyl-sulfate polyacrylamide gel electrophoresis (SDS-PAGE) analysis [37]. The digital images captured for the unmodified (i.e., no PEG and/or antibody) and PEG/antibody-modified Au-SPEs showed a change in the color of the modified electrode surface (**Figure 1b**). In addition, the Au-SPEs were digested using tris (2-carboxyethyl) phosphine (TCEP), and the UV-Vis spectrophotometry technique was used to confirm the presence of antibodies in the eluted solution [38-40]. The characteristic absorbance peak for PEG was observed near 220 nm and the absorbance peak for the antibody was observed near 260 nm, compared with unmodified control Au-SPEs (**Figure 2c**). To further characterize the anti-KPC2 IgG antibody-modified Au-SPEs, a horse radish peroxidase (HRP)-based colorimetry technique was used. The unmodified and anti-KPC2 IgG-modified electrodes were incubated with HRP-conjugated anti-IgG for 15 minutes, and then it was gently washed three times, then 10 µL of 3,3’,5,5’-tetramethylbenzidine (TMB) substrate was added on to the Au-SPEs (**Figure 2a**). The complex was incubated with TMB for 10 minutes and a strong blue color was seen in the antibody-modified SPE. This suggests that the HRP-bound anti-IgG was able to successfully bind the anti-KPC2 antibody and was able to reduce the TMB to produce a blue color. The TMB added to the unmodified SPE did not produce any color change indicating no presence of anti-KPC2-antibody. Further, the reduced TMB substrate was collected and the absorbance for the formed complex was measured at 652 nm. The sharp rise in the absorbance at 652 nm confirmed the presence of anti-KPC2 antibodies on the surface of Au-SPE and its labeling with HRP. The KPC2 antibody-modified Au-SPEs were also incubated with protein-G coupled with fluorescein isothiocyanate (FITC) for 15 minutes (**Figure 3 a**). Protein G tends to bind to most IgG molecules at near physiological pH at room temperature (25 °C). The bound antibody complex was then digested using TCEP from both unmodified and modified SPEs, to release the IgG-protein G-FITC complex from the surface of Au-SPE, and the fluorescence signal was measured for the released complex at 528 nm. The coupled FITC to protein G was able to produce a fluorescent signal at 528 nm, which was ∼68.2% higher than control sample of unmodified electrodes, indicating a successful conjugation of antibody to the surface of Au-SPE. To test the efficiency of the antibody-modified SPEs to capture the target KPC2 target enzyme, antibody-modified Au-SPEs were used to capture the KPC2 enzyme (at a very low concentration of 20 ng/mL). The SPEs were incubated with the KPC2 enzyme for 30 minutes and washed 3 times using 10 mM phosphate buffer pH 7.2 to remove excess unbound enzyme. The complex was then digested using TCEP and the released complex was used to perform SDS-PAGE and FTIR analyses. FTIR analysis results showed absorption bands of amide I group at 1625 cm^-1^ (C−O stretching vibration of peptide linkages), and COOH group (C=O stretching vibration) at around 1784.05 cm^-1^, providing proof of the successful surface modification of the beads (**Figure 4a**), indicating a successful surface-modification with antibody and enzyme capture using the modified Au-SPEs [41-43]. The TCEP digested complex from electrodes’ surface was also analyzed using SDS-PAGE technique. The presence of band specific to KPC2 antigen at 27.2 KDa in the TCEP digested sample indicates successful and effective capture of the KPC2 enzyme by the antibody coated on surface of Au-SPE. In addition, we used UV-Vis spectroscopy to compare the capture efficiency of KPC2 enzyme on the surface of Au-SPE modified using the developed PEG protocol with the traditional NHS-based chemistry that relies on using 3-(2-Pyridyldithio)propionic acid *N*-hydroxysuccinimide ester (SPDP). The NHS ester reacts with the amine groups of anti-KPC2 IgG antibodies, and the pyridyldithiol reactive groups in the generated SPDP-modified antibody molecules bind to the surface of Au-SPEs. The results indicated 12.6 ± 2.1 folds increase in the capture of the target KPC2 enzyme using our PEG-based site-specific direct conjugation approach (**Figure 4c**).

**Figure 2.**
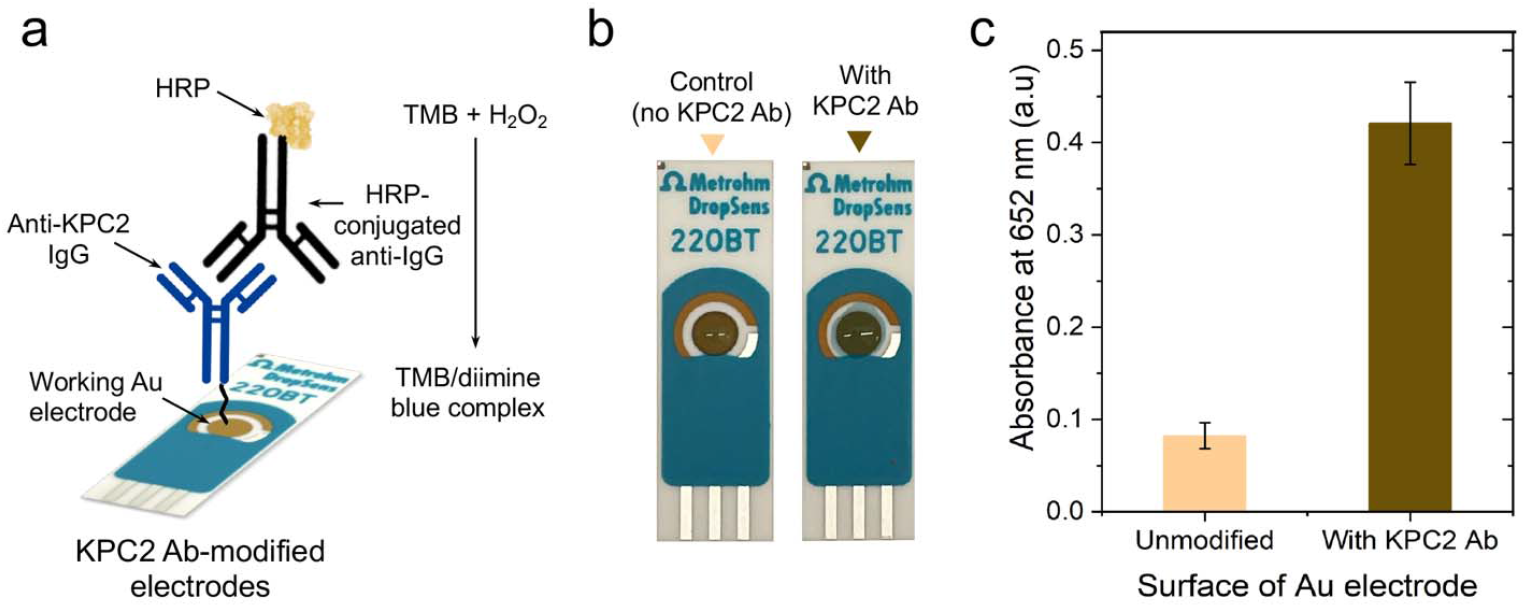
Characterization of antibody-modified Au-SPEs using HRP-based colorimetry. (a) Schematic presentation of the HRP-based colorimetry used to characterize the surface of the modified electrodes. The unmodified and modified Au-SPEs were incubated with HRP-conjugated anti-IgG and washed. Aliquots of TMB (10 µL) substrate were added to each strip and incubated for 10 minutes. (b) Digital images captured for the strips after TMB addition, showing a color change of TMB suggesting the presence of HRP. (c) The UV-Vis absorbance of TMB collected from the surface of electrodes was measured at 652 nm.

**Figure 3.**
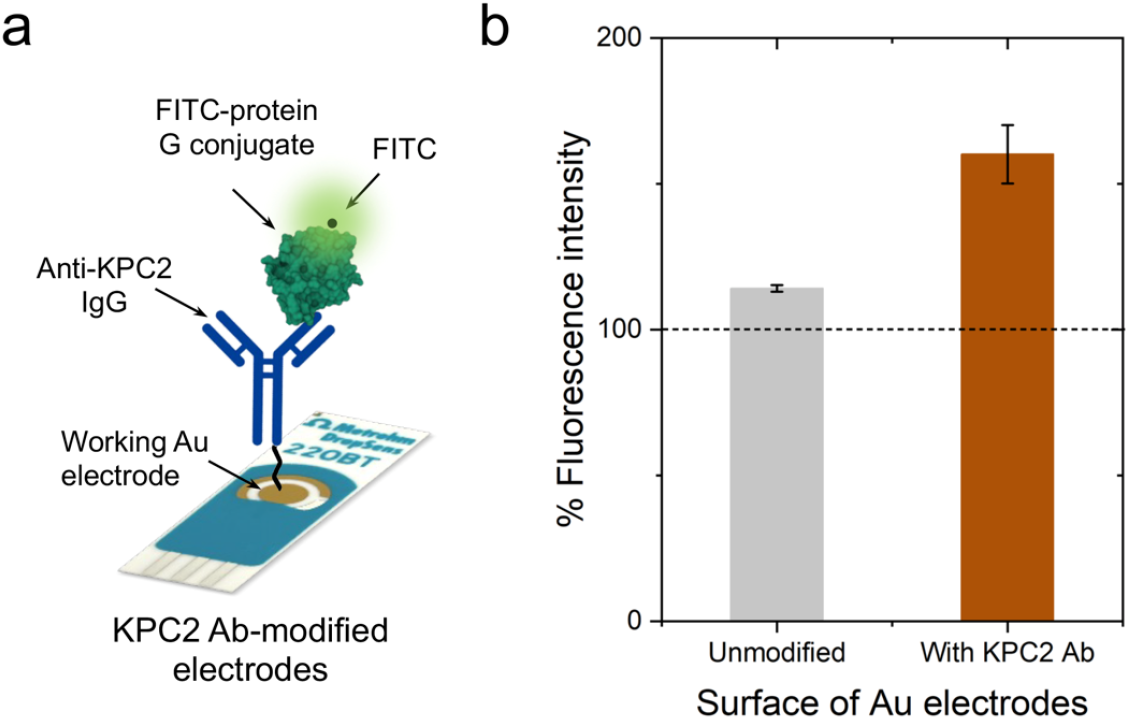
Characterization of antibody-modified Au-SPEs using FITC-based fluorometry. (a) Schematic presentation of the FITC-based staining protocol for Au-SPE characterization. The antibody-coated Au-SPEs were incubated with protein G coupled with FITC, which reacts with the antibody present on the surface of the electrode. (b) The surface-modified and unmodified electrodes were digested with TCEP to release the coupled antibody stained with protein G-FITC. Fluorescence intensity was measured for the released analyte at 528 nm.

**Figure 4.**
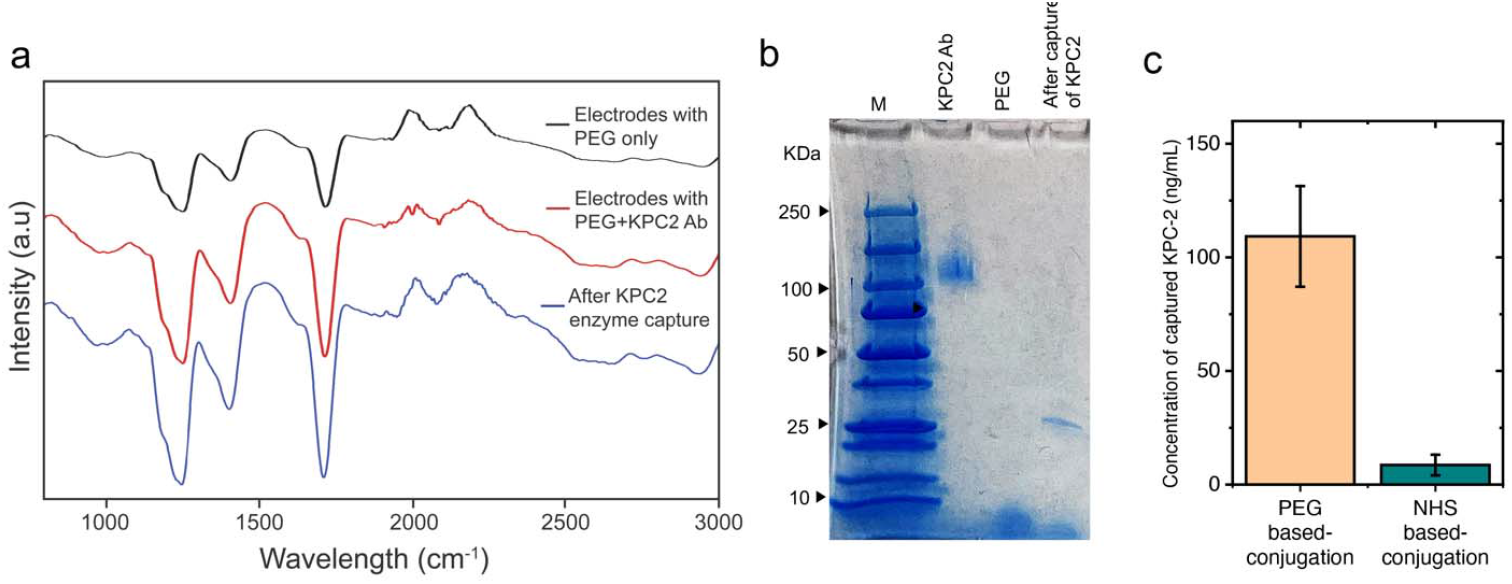
Characterization of antibody-modified Au-SPEs and KPC2 enzyme capture efficiency. (a) FTIR analysis of the modified (in red) and unmodified electrodes (in black), and modified electrodes after enzyme capture (in blue) (protein specific range at what wavenumber?) post TCEP digestion (b) SDS-PAGE analysis of modified with anti-KPC2, unmodified and modified with anti-KPC2 with enzyme captured electrodes after TCEP digestion, showing the presence of bands specific to antigen at 27.2 KDa. (c) The concentration of KPC2 enzyme captured on the surface of Au-SPEs modified with KPC2 antibodies using the developed PEG-based conjugation compared with the traditional NHS-based conjugation chemistry. The surface of Au-SPEs (5 electrodes) was modified with antibody and loaded with 10 µL of KPC2 enzyme at a concentration of 20 ng/mL and incubated for 30 minutes at room temperature. After incubation, the unbound target enzyme was collected and the concentration of the captured target KPC2 enzyme was estimated from the total concentration of the loaded KPC2 enzyme using UV-Vis spectroscopy.

In this study, we presented a novel conjugation chemistry for developing enhanced KPC2 enzyme testing at POC settings using SPEs-based electrochemical immunoassay [44]. Our approach relied on a site-directed reaction that used a heterobifunctional PEG polymer to link oxidized polyclonal anti-KPC2 antibodies to the surface of Au-SPEs. We achieved PEGylation of the Au-SPE using SH-PEG-hydrazide, which left the hydrazide groups free to react with the oxidized Fc regions of anti-KPC2 polyclonal antibodies. Our results demonstrated that this site-directed conjugation approach offered a viable alternative to traditional NHS-based chemistry for the modification of Au-SPEs with antibodies. By using directional conjugation, we were able to obtain highly efficient and specific binding of the antibody to the enzyme, which is crucial for designing and developing accurate detection of KPC2. We also conducted a series of tests to confirm the presence of the antibody on the surface of the modified Au-SPEs, including colorimetry, fluorometry, UV-Vis and FTIR spectroscopy, and SDS-PAGE analysis. Our findings demonstrated the successful capture of the KPC2 enzyme using the modified Au-SPEs, with the antibody-modified Au-SPEs capturing the enzyme at a very low concentration of 20 ng/mL. These promising results may have implications for the development of diagnostic tools for the early detection of infections caused by KPC2-producing bacteria, which is crucial for effective management and control of antibiotic-resistant infections. Overall, our study highlights the potential of our PEG-based site-directed conjugation approach for the development of sensitive and specific POC diagnostics for a range of diseases and conditions.

## Methods

### Au-SPE surface activation with PEG

The surface of Au-SPEs is cleaned using 75% ethanol solution for 5 minutes. The cleaned electrodes were washed with Milli-Q water and allowed to dry at room temperature (25 °C). Freshly prepared PEG solution (20 mg/mL; thiol-PEG-hydrazide) was drop-casted on the surface of the working electrode of Au-SPE, and incubated for 1 hour. After the reaction was completed, the modified electrodes were washed with water 3 times and used for antibody coupling reaction.

### Antibody oxidation and coupling to Au-SPE

Aliquots of 20 µL of antibody (1.2 mg/mL) were mixed with 10 mM of sodium metaperiodate (pH = 5.5) acidified with 0.1 M sodium acetate buffer (pH = 5.5), and incubated at 4 ºC in dark for 20 minutes. Post-oxidation, the antibody was washed with phosphate buffer (pH=7.4) using centrifugal filter units with a cut off value of 50 kDa. Oxidized antibody solution was incubated with the PEG-modified Au-SPE.

### Characterization of antibody-modified electrodes

The surface modification of Au-SPEs was characterized by using UV-Vis spectroscopy. The modified and unmodified SPEs were digested with TCEP (three to five SPEs per test), and the protein concentration in the eluted solution was measured using NanoDrop 2000/2000c Spectrophotometer (Thermo Fisher Scientific, Inc., USA). To perform the HRP-based colorimetric assay, the antibody-modified and unmodified SPEs were incubated with HRP-conjugated anti-IgG antibodies for 20 minutes and washed 3 times gently. 10 µL of TMB substrate was added to each electrode and incubated for 10 minutes. A blue color was developed after the incubation period. The absorbance intensity of the formed blue color was also measured at 652 nm. The unmodified and modified SPEs were incubated with protein G coupled with FITC to bind the IgG on the surface of the electrode. After 15 minutes of incubation, the SPEs were washed thrice gently to remove excess unbound protein G. The electrode surface was then digested with TCEP to release the bound protein G with FITC, and fluorescence intensity of the released solution was measured at 528 nm to confirm the binding of Protein G to the antibodies on the surface of electrodes.

### Enzyme capture assay and assessment of enzyme capture efficiency

We incubated antibody-coupled SPEs and PEG-modified SPEs (no antibody) with the target KPC enzyme (10 mg/mL). The surface of each electrode was washed surface were treated with 100 mg/mL TCEP solution to release the bound protein. UV-Vis and Fourier transform-infrared (FTIR) spectroscopy techniques were performed to confirm the enzyme capture.

### Ultraviolet-visible (UV-Vis) spectroscopy

Absorption spectra were measured on NanoDrop 2000/2000c Spectrophotometer (Thermo Fisher Scientific, Inc., USA). Concentration was calculated by measuring the absorbance of 1□µL of the sample at 260/280□nm at room temperature. Each sample was tested 3-5 times, and average values are presented.

### Fourier transform infrared (FTIR) spectroscopy

FTIR spectra in the region of 2000-500 cm^-1^ were collected in absorbance mode with a FTS 135 BIO-RAD FTIR spectrometer.

### SDS gel electrophoresis

Protein testing was performed using SDS-PAGE on 20% Tris-Glycine gel. The collected KPC2 antibody and captured enzyme aliquots were digested in digesting buffer and heated for five minutes at 95 ºC on a heat block. For gel electrophoresis analysis, 12 μL of a protein standard and 35 μL of the centrifuged samples were loaded on the gel using a mannoprotein electrophoresis apparatus (Bio-Rad, Hercules, CA). The samples were electrophoresed under 90 V for 50 minutes. After electrophoresis was done, the gel was rinsed in water for 3 minutes. Then the gel was stained in the Biosafe Coomassie blue stain for about 1 h. Finally, the gel was de-stained and photographed.

## Data Availability

All data produced in the present work are contained in the manuscript

